# Developing pediatric eye care program in Nepal: Insurmountable Mountains can be reduced to mole hills. The Orbis experience in Nepal

**DOI:** 10.1101/2023.06.19.23291618

**Authors:** Rishi Raj Bora, Parikshit Gogate, Saibaba Saravanan, Sailesh Kumar Misra, Reeta Gurung, Yuddha Dhoj Sapkota, Srijana Adhikari, Purushottam Joshi, Suresh Raj Pant, Kabindra Bachracharya, Hari Bikram Adhikari, Govind Gurung, Sanjay Kumar Singh

## Abstract

**Background:** Nepal straddles the Himalayas and its geography has posed challenge to delivering eye care in children. This manuscript describes the pediatric eye care facilities developed, and children examined and treated under the Orbis International’s childhood blindness initiative.

**Methods:** Between 2010-2017 Orbis International had supported the Nepal Netra Jyoti Sangh (NNJS) and Tilganga Institute of Ophthalmology (TIO) to develop pediatric eye care centers in major hospitals, along with their outreach. This manuscript looks at the impact of that intervention. Reports of Nepal Netra Jyoti Sangh, Tilganga Institute of Ophthalmology and Orbis International were studied, along with publications on ophthalmology from Nepal. Eight child friendly pediatric eye care centers were set-up, pediatric eye care teams were trained, an outreach program especially for children’s eye problems was designed and outcome monitoring and research promoted.

**Results:** Between 2010–17 1,281,153 children had been examined by 8 pediatric eye centres (average 20,018 per hospital per year, range 10,729 –39,485) and the average outpatient per month per hospital was 1668 (range 894 - 3,290). Between 2010 – 2017, 42,430 children had been operated upon by 8 hospitals (average 663 per hospital per year, ranged from 96 – 1,465) and the average pediatric eye surgery per month per hospital was 55 (ranged from 8 to 122).

In the years 2018 & 2019 (21 months) post-project, all the 8 hospitals had operated on 14,252 children (average of 1,782 per hospital, range 185 - 4438) and the average pediatric eye surgery per month per hospital was 85 (range 9 to 211). 19 publications on pediatric eye care were published in indexed journals.

**Conclusion:** Orbis childhood blindness amelioration initiative in Nepal resulted in more than a million children examined, and >50,000 children underwent eye surgeries in the 8 pediatric eye care centres and the good service continued after the project was over.

## Introduction

Nepal was the world’s only Hindu kingdom that became a secular republic in 2008 after a decade-long Maoist insurgency. Nestled in the mighty Himalaya mountains, it has one of the most dramatic landscapes, from near sea level to over 29000 feet within a few hundred kilometers. One of the poorer parts of Asia, due to its terrain and remote landlocked location, it was also wracked by internal strife. [1] Nepal is mainly divided into three physiographic areas: Mountain, Hill and Terai. These ecological belts run east-west and are vertically intersected by Nepal’s major, north to south flowing river systems. Himalayan Region – 15 per cent of the country’s total land is covered with snow-capped mountains in the northern part, and altitude ranges from 4,877 to 8,848 meters, including the world’s highest peak Mount Everest (8,848m). Hilly Region – the hills and mountains cover 68 per cent of the country’s total land in the central part of Nepal. The altitude varies from 610 to 4,877 meters in this region. Terai Region-In the southern part, the plain area of the Terai region covers 17 per cent of the country’s total land but has nearly half the population. Nepal’s gross domestic product (GDP) for 2021 was estimated at over $36 billion. Nepal has had a steady improvement in per capita income and health care indices in the past four decades. [1,2] Nepal Netra Jyoti Sangh (NNJS) an umbrella non-governmental organization for providing eye care under the aegis of Government of Nepal had helped set up large hospitals in each zone of the country. [3] Most service was for adults, but children too needed care, as 40% of populace was <18 years old. A large prevalence of blindness survey was conducted in Nepal in 1980s. [4]. A total of 17,423 children aged 0-15 were examined during the National Blindness Survey in 1981 and the prevalence of bilateral blindness in 10-19 year olds was 0.14 per hundred. [4]

Orbis is a non-aligned, non-profit global development organization whose mission is to preserve and restore sight by strengthening the capacity of local partners in their efforts to prevent and treat blindness. [5] Through the Childhood Blindness Initiative, Orbis is working with not-for-profit organizations in the eye care sector to establish well-equipped Children’s Eye Centers across the globe with a particular emphasis on under-served rural areas. It aims to provide access to quality eye care to millions of children in each of the country it works by setting up pediatric eye care centres with its partners; and once established and operational, these child eye care centers shall continue to serve future generations. Till the year 2000, blindness prevention interventions in Nepal had been cataract centric and the whole country did not have a single dedicated pediatric eye care center to treat children. In July 2004, under ORBIS India program, Shree Rana Ambika Shah Eye Hospital (SRASEH) at Bhairawa near Lumbini (Buddha’s birthplace) in Terai region, one of the high-volume eye hospitals of Nepal, was identified for establishing a dedicated child eye care center. Currently it is named as Lumbini Eye Institute and Research Center (LEI).

ORBIS had commissioned a national survey in Nepal in the year 2007 to identify gaps in current availability of infrastructure and trained human resources related to pediatric eye care services. As per the survey, there were a total of 6 trained pediatric ophthalmologists in Nepal, one trained pediatric ophthalmologist for every 4.3 million population. The program was then extended to establish seven more pediatric eye centres in Nepal.

There have been few studies that have documented the change in health care delivery status in developing countries. Thomas et al (2005) provided a review of eye care in India.[6] Durkin (2008) reviewed the literature available on eye health care programs in Australia for aboriginals.[7] Faal et al reported the prevalence of blindness in Gambia, a small West African country after a ten year intervention in terms of a national eye care program.[8] Aghaji et al (2018) described the strengths, challenges and opportunities of implementing primary eye care in Nigeria.[9] There have been no such publications documenting eye care from Nepal.

This manuscript reports the results and outcomes of the decade plus interventions aimed to improve eye care service delivery in children in Nepal from 2004 to 2019, during the project period and after completion, for sustainability.

## Methods

Permission was sought and obtained from the ethics committee of Nepal Netra Jyoti Sangh, Kathmandu, Nepal. Annual reports of Orbis International, India Country Office, Nepal Netra Jyoti Sangh and Tilganga Eye Institute (later renamed Tilganga Institute of Ophthalmology) were studied as were the project reports, quarterly reports and midterm and final evaluation reports. No individual medical records, either of adults or children, were directly accessed for this study. Orbis International, India Country office had initially supported Shri Rana Ambika Shah Eye Hospital (SRASEH, later called Lumbini Eye Institute & Research Center) in Bhairava, Lumbini in the Terai region of Nepal, from 2004-2008. Orbis helped refurbish the outpatient department, train human resource, purchase equipment and shore up outreach activities.

Orbis first trained a pediatric eye care team, an ophthalmologist, optometrist, anesthetist, orthoptist, pediatric oriented nurse and counselors. [10] They were existing team members who were trained for pediatric orientation at a center of excellence in India (Aravind Eye Care System, Madurai, Tamil Nadu). A paramedic and/ or technician was trained in basic equipment maintenance at the same institute. This was supplemented by short training programs abroad the Orbis Flying Eye Hospital which visited Kathmandu and hospital based programs (HBPs) where volunteer faculty visited, stayed and taught trainees in their own hospitals for a week.

A child friendly pediatric eye care center was developed with a dedicated children’s play area, breast feeding area, toilets for children and a pediatric ward. [10] Facilities were created for general anesthesia in the operating rooms. The management and team of the eye hospital were trained in gender sensitization, and to ensure equal access and care to patients irrespective of their gender, ethnicity and socioeconomic status.

The strategies to improve pediatric eye care used first in the SRASEH, Lumbini region from 2004-2007 were followed by six other hospitals operated by the Nepal Netra Jyoti Sangh (NNJS) from 2010-2017. They were 1. Himalaya Eye Hospital (HEH), Pokhara; 2. Sagarmatha Chaudhary Eye Hospital (SCEH), Lahan; 3. R M Kedia Eye Hospital (KEH), Birgunj; 4. Mechi Eye Hospital (MEH), Birtamode; 5. Geta Eye Hospital (GEH), Dhangadi; 6. Fateh-Bal Eye Hospital (FBEH), Nepalgunj and the Tilganga Eye Institute in Kathmandu, the national capital. Together they covered all the five zones of Nepal. Figure 1 shows the map of Nepal with its 5 zones and the location of the hospitals in 2007. Figure 2 shows the map of Nepal with the newly constituted 7 provinces and the location of the eight pediatric eye care centres in 2019.

**Figure 1:**
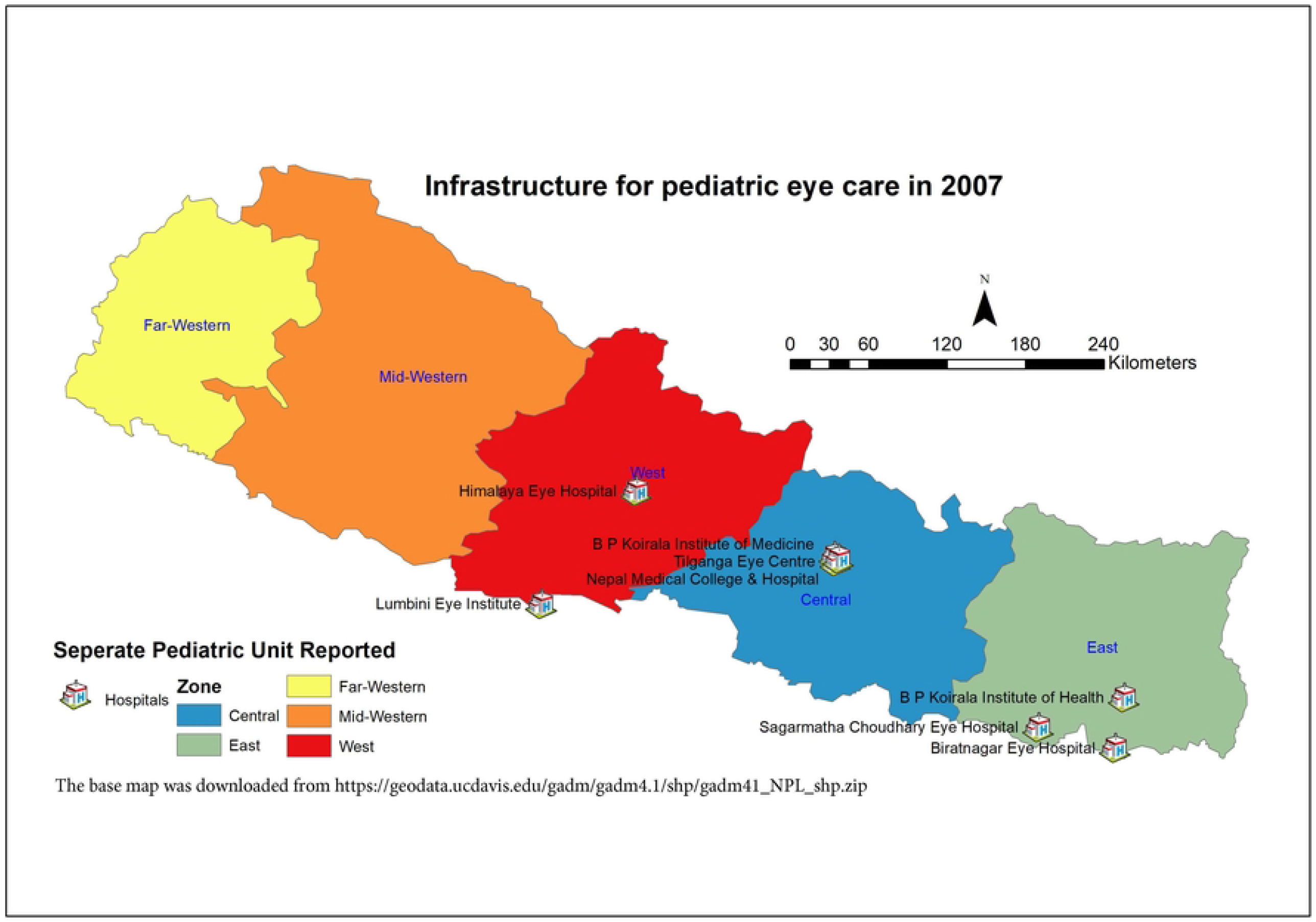
Map of Nepal showing the 5 zones and location of eye centers in 2007. This map was created using freely available country boundary base map GADM.org, rendered in ArcGIS. The base map was downloaded from https://geodata.ucdavis.edu/gadm/gadm4.1/shp/gadm41_NPL_shp.zip under a CC BY license

**Figure 2:**
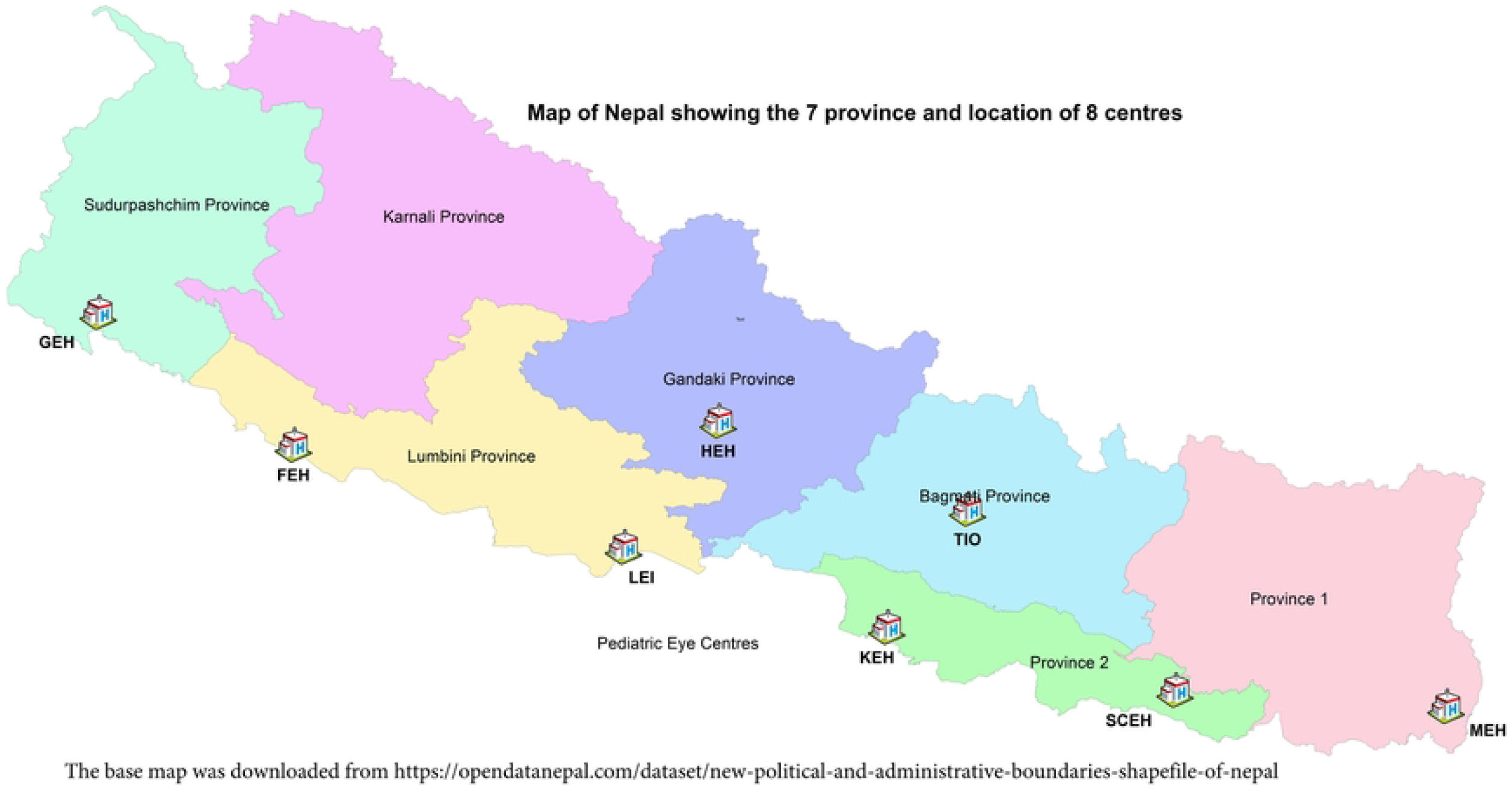
Map of Nepal showing the 7 Province and location of 8 centers in 2019. This map was created using freely available country boundary base map opendatanepal.com, rendered in ArcGIS. The base map was downloaded from https://opendatanepal.com/dataset/new-political-and-administrative-boundaries-shapefile-of-nepal under a CC BY license.

Primary eye care centers of the hospital were strengthened by donating pediatric visual acuity testing equipment, emphasizing school eye screening and developing pediatric outpatient facilities. [10] The REACH project helped strengthen the outreach by examining school children by teaming up with the primary eye clinics attached to the hospitals. [11]

The network of 106 primary eye care centers working under aegis of NNJS, located in the 77 districts of Nepal added as a feeder to the secondary eye centers and helped screen children with visual impairment and direct them to secondary and tertiary centers where general anesthesia facilities were made available. 28 such primary centres were upgraded for delivering pediatric eye care. Partner hospitals were encouraged to use electronic medical record systems and measure the outcome of pediatric eye surgeries performed. Numerous meetings, conferences and hospital based programs were conducted.

The project encouraged the hospitals to have regular follow-up after paediatric ocular surgery. Regular follow-up by itself improves outcome as shown by a publication from Nepal in JAAPOS and the Orbis supported Miraj paediatric cataract study in Maharashtra, India.[12,13] The project made measuring outcomes mandatory and ensured documentation of follow-ups. The teams were encouraged to conduct clinical and operational research. The scientific publications that ensured from the data were also documented.

The project also had a health promotion and education component. Efforts were made by Orbis to raise awareness on eye health among parents and care givers: Counsellors in Orbis partner hospitals were trained to strengthen counselling of children and parents visiting the hospitals. The Training Center for pediatric ophthalmology established at Lumbini Eye Institute had a refresher course for counsellors to improve counselling in children’s eye care. The teams used the concept of *Bhaat Bhahdur – Saag Bahadur* (**भात बहादुर** - **साग बहादुर**) in patient communication, especially for children in the Children’s Eye Centers for raising awareness among children and parents visiting the center. Bhaat is rice, Saag is green leafy vegetables, Bahadur means brave and is a common gorkha-Nepali name amongst boys. Children who subsist only on rice were more likely to be Vitamin A deficient and malnourished, as compared to those who consume green leafy vegetables too. The project developed relevant IEC materials (brochure, leaflets and posters) on child eye health and general eye health awareness and distributed these in the community during outreach activities and through the fixed facilities (Primary eye Care Centers and Hospitals). Biratnagar Eye Hospital has created dedicated infrastructure for patient counselling and education in the hospital to provide adequate emphasis on patient counselling and systematize it.

The data was collected in excel software (Microsoft Inc.).

## Results

Lumbini Eye Institute, the first hospital to set up a dedicated pediatric eye unit was examining 5,189 children before Orbis intervention in the year 2003 (432 per month) and with the Orbis support had improved their capacity to examine about 15,556 children by the year 2007 (1296 per month). The overall children examined between 2004-2007 were 51,887 (avg 12,972 per year). The other 7 hospitals had a pediatric eye care centre in 2010. Table 1 shows the geographic coverage by each partner hospital.

**Table 1:**
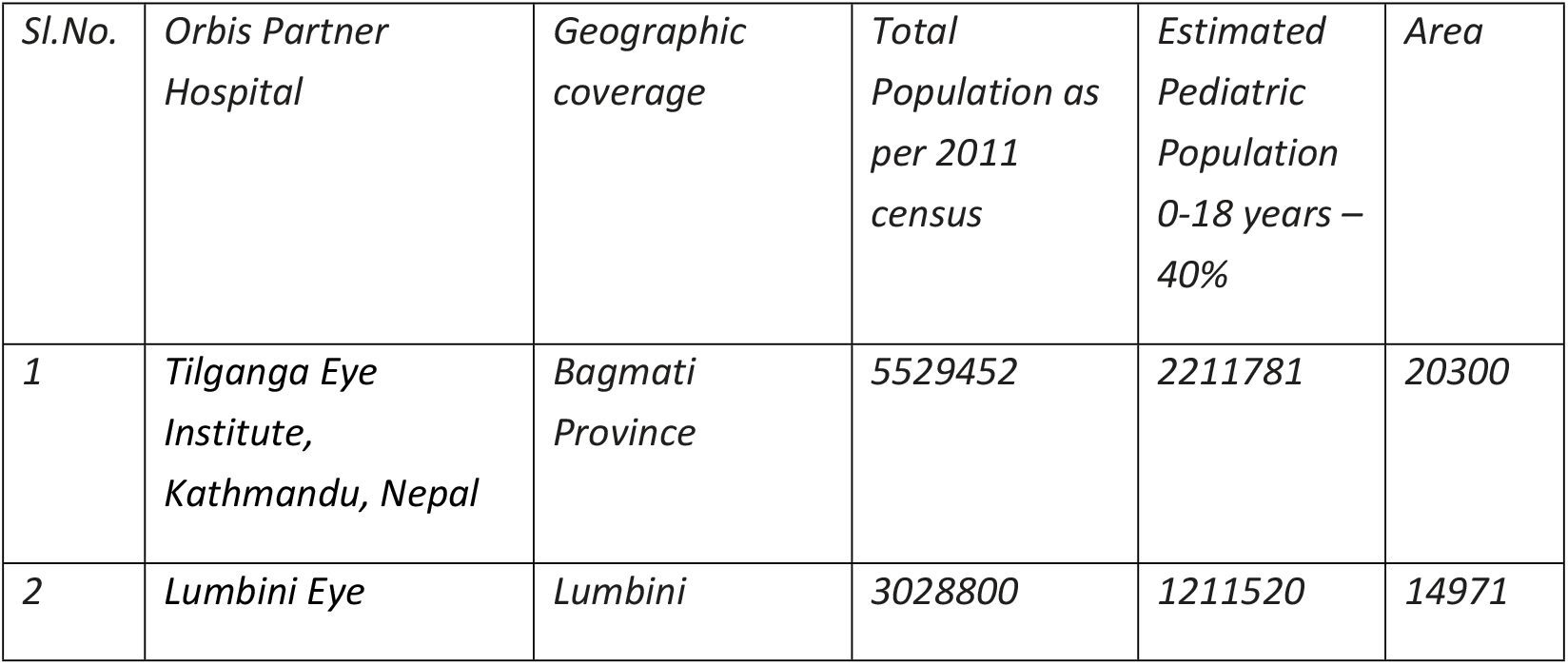
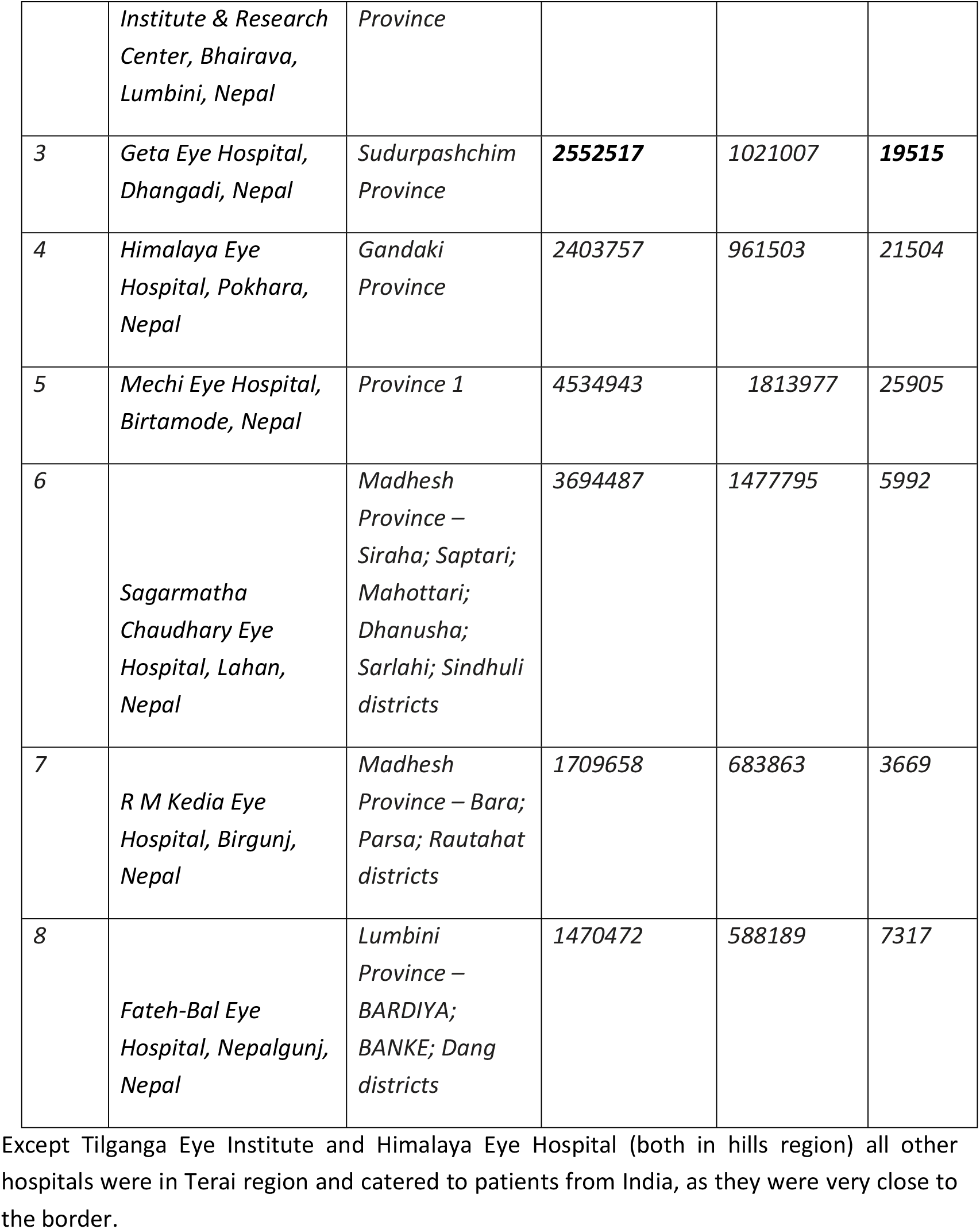
shows the geographic coverage, population and area covered by each hospital.

Table 2 shows the out-patient children visits and pediatric eye surgeries before (2010), during (2010-17) and after (2018-19) the project. The 2018-19 figures are for 21 months.

**Table 2:** Overall pediatric eye clinic out-patient visits and pediatric eye surgeries before, during and after the project period.

| **8 centres** | **2010** | **2010-2017** | **2018-19** |
| --- | --- | --- | --- |
| Out-patient children (Total) | 94,114 | 12,81,153 | 3,66,377 |
| Out-patient children (Average) per year per hospital | 11,764 | 20,018 | 26,170 |
| Out-patient children (Range) | 0-28,419 | 10,729-39,485 | 13,558-47,737 |
| Outpatient children per month per hospital (average) | 980 | 1668 | 2181 |
| Outpatient children per month per hospital (range) | 0-2,368 | 894-3,290 | 1,130-3,978 |
| Paediatric eye surgeries (Total) | 2715 | 42430 | 14252 |
| Paediatric eye surgeries (Average) | 339 | 663 | 1782 |
| Paediatric eye surgeries (Average) | 0-1019 | 96-1465 | 185-4438 |
| Pediatric eye surgeries per month per hospital (average) | 28 | 55 | 85 |
| Pediatric eye surgeries per month per hospital (range) | 0-91 | 8-122 | 9-211 |

The number of paediatric surgeries performed had more than doubled on average. Among the total pediatric surgeries performed, cataract was the highest (42%, ranging from 24% - 62%), followed by oculoplastic/ptosis/lid surgery (11%, ranging from 0.3% to 25%) and corneal tear repair/transplant (10%, ranging from 7% to 17%).

The different types of surgeries performed between 2010-2019 by all hospital are shown in the following Table 3.

**Table 3:** Types of pediatric eye surgeries performed

|  |  |
| --- | --- |
| Cataract | **41.8%** |
| Strabismus/Squint | **2.9%** |
| Pterygium | **0.5%** |
| DCR/DCT | **1.5%** |
| Glaucoma | **1.6%** |
| Cornea Repair/Transplant | **10.2%** |
| Oculoplastic/Ptosis/Lid Surgery | **10.9%** |
| Vitreous/Retina | **8.4%** |
| Laser | **0.1%** |
| Others | **22.5%** |
| Total Children Operated | **100%** |

Others include lid tear repair, conjunctival tear repair, corneal foreign body removal, probing and syringing for nasolacrimal duct obstruction, detailed evaluation of peripheral retinal under general anesthesia. They were included under ‘surgeries’ as they were performed in the operation theatre with requisite consent.

There were hardly any general anaesthesia and paediatric eye surgery facilities in most large eye hospitals before the Orbis childhood blindness project. In 2004 at LEI and also in 2007 at the 7 hospitals, ketamine anaesthesia was being used, which was not the best preferred technique both for open globe surgeries and for strabismus surgeries. With the development of clinical protocols, training of staff, anaesthesiology support had improved and safe anaesthetic agents were used now in the 8 hospitals. Intubation inhalation anesthesia service and use of laryngeal masks for paediatric general anaesthesia had become common. Surgery on infants (age<1 year) had been started.

The 2007 study had indicated the lack of pediatric teams available in the hospitals and the available resources were unevenly distributed, with most services concentrated in the big urban areas and the capital city of Kathmandu. In 2007, there were a total of 6 trained paediatric ophthalmologists in Nepal. Out of the 6, one was trained under Orbis project from LEI. There were no paediatric oriented ophthalmologists in the country, though many other ophthalmologists regularly attended to paediatric patients. Figure 3 shows the pediatric eye care teams available in Nepal.

**Figure 3:**
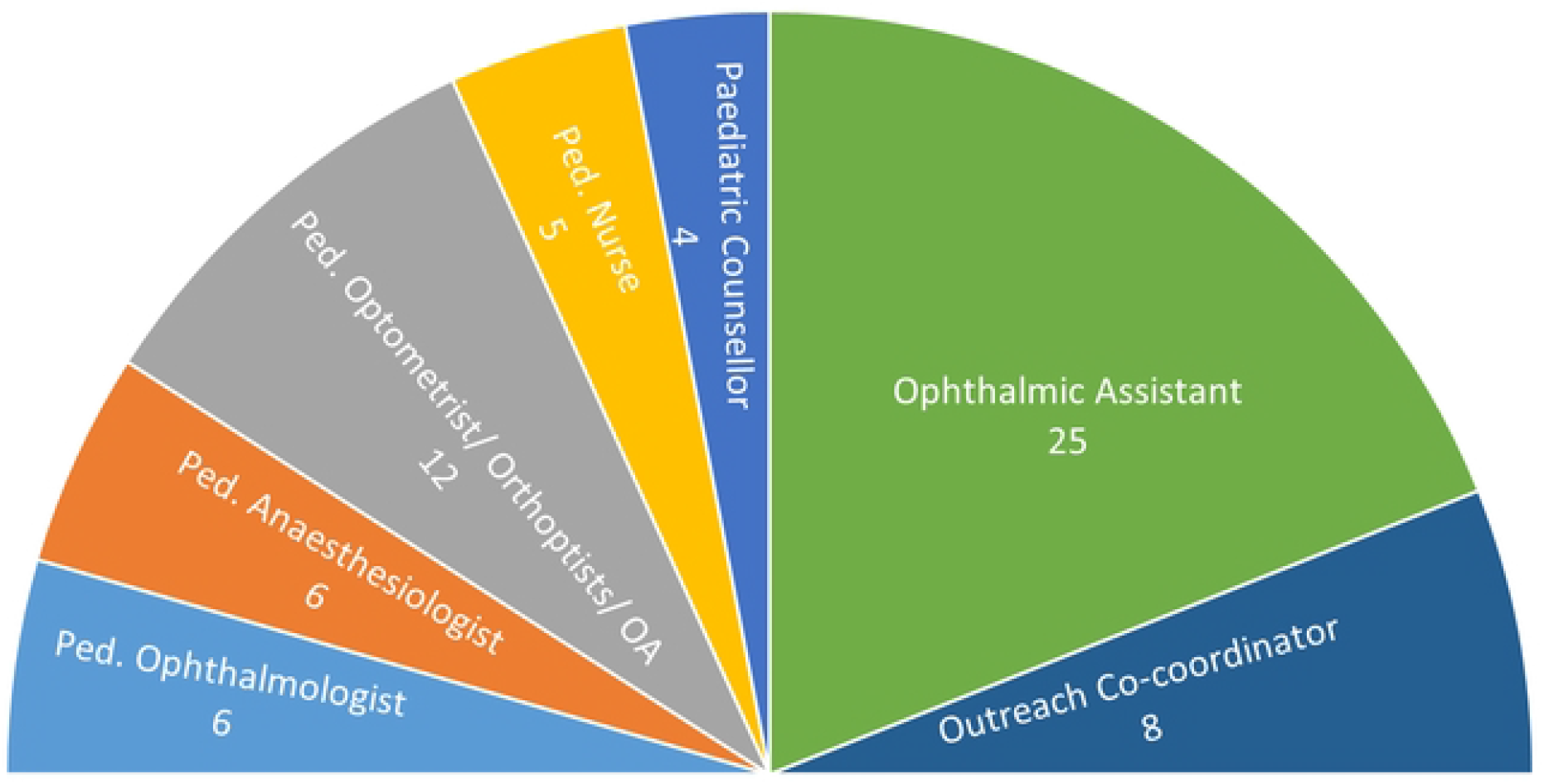
Pediatric teams available in Nepal - 2007 Nepal HR Report

As of 2019, it is estimated that there were about 22 paediatric ophthalmologists in the country and 12 of them were at Orbis partner facility, and of these 7 had been trained by Orbis support (6 in India and 1 in LEI, Nepal); and in addition 6 ophthalmologist have been trained at LEI. Of the 22, 18 paediatric ophthalmologists were either directly trained, serve at partner facility or trained at Orbis supported training facility. In addition, more than 25 ophthalmologists were trained under the flying eye hospital program and hospital based programs that were part of the project to develop human resource.

In 2007, the country had 5 paediatric nurses (1 trained by Orbis); 4 Paediatric counsellors (1 trained by Orbis) and 12 Ped. Optometrist/Orthoptist (3 trained by Orbis). By 2017, in addition to the ophthalmologists, various other staffs were trained in paediatric eye care were 18 optometrist/ Orthoptists, 19 paediatric nurses, 15 paediatric counsellors, 11 ophthalmic assistants in outreach programs, 9 administrators/project manager and 5 bio-medical technicians. In addition, 13 ophthalmic assistants were trained under the flying eye hospital program and hospital based programs.

By 2019, except one hospital, all the hospitals had full-time paediatric ophthalmologists and had trained optometrists/orthoptists, paediatric nurses and counsellors. Even though few trained staff had left the organization, all the hospitals tried to maintain the paediatric team composition.

In 2007, there were no anesthetists oriented with ophthalmic pediatric surgery. However, 6 anesthesiologists and 6 anesthesia assistants/technicians had been trained through the flying eye hospital program and Hospital-Based Programs (HBPs). Due to the high demand for anesthetists, all the hospitals excluding those in Kathmandu, were not able to get anesthetists every day. Very small children and infants were rarely taken for surgery earlier, but this changed during the project period.

At the beginning of 2018, LEI had organized the first national paediatric ophthalmology and strabismus conference, where most of the paediatric and paediatric oriented ophthalmologists in the country participated. SCEH and TIO in 2016 had started providing a one-year fellowship program owing to the rise in demand for paediatric ophthalmologists and the interest the project has created among the ophthalmologists. Currently, about five paediatric ophthalmologists are being trained each year in Nepal.

All these centres pursue research in childhood blindness and have notable publications in various journals. Manuscripts on children’s eye health in Nepal were published in pubmed indexed journals. [10, 14-28] The hospital teams had publications on low vison services for children [14-17], refractive errors [18, 19] and ocular injuries in children [21, 22].

All the centres now have optometrist training programs and orthoptics is part of the curriculum. Hands-on training in pediaric eye care is imparted to the budding optometrists.

The dedicated pediatric ophthalmology units had the highest OPD load amongst all ophthalmic subspecialties like cornea, retina, glaucoma and oculoplastic, again reaffirming the fact that there was a great need for setting up such a service. Hospitals too had set up a dedicated low vision service.

At LEI, during the years 2004-2007, Nepalese children accounted for 59% of the OPD and 40% of the surgeries. About 41% of OPD patients and 60% of the surgeries were Indian children whose parents had got them from across the border for treatment. Between 2010 and 2019, paediatric referral had increased from remote and rural areas of Nepal and the 8 hospitals had covered the entire Nepal (77 districts) and 9 states of India Uttarkhand, Uttar Pradesh, Bihar, West Bengal, Jharkhand, Assam, Meghalaya, Arunachal Pradesh, Nagaland) in terms of service delivery. By 2019, 76% of the paediatric OPD and 56% of paediatric surgery were of the native Nepalese population. This had increased from 30% in 2010. 24% of the outpatient and 44% of the paediatric eye surgery in the 7 out of 8 (Excluding HEH) hospitals in the Terai region mostly came from the neighbouring Indian states of Uttar Pradesh, Bihar and West Bengal.

The above data was only from the base hospital’s pediatric eye care centres. More children had been examined at the 28 upgraded Primary Eye Care Centres/ Child Eye care Centres. In 2010, in these centers children accounted for only 2% of the total out-patients which increased to >10% in 2019. Children were also referred from outreach camps, school screening program and through Female Community Health Volunteers (FCHV).

## Discussion

The intervention by Orbis with NNJS and TEI had greatly increased the volume and scope of pediatric eye care services in Nepal and also benefitted the surrounding states of India. In 2004-8 Orbis International first supported the implementation of the pediatric eye care center and project in Lumbini. Its success led to the project with 6 other NNJS Eye Hospitals and the Tilganga Institute of Ophthalmology. More importantly, the volume of service rendered during the project duration of 2010-17 was maintained in 2018-19, after the project was over. Cataract and eye injuries in children were managed well. The increased number of primary and secondary eye centers providing eye care for children, coupled with increased school enrollment ratio and better roads, reduced the barriers of awareness (lack of) and distance. There was increased awareness about eye problems in children which improved the demand for services. Pediatric ophthalmology had developed as a sub-specialty and pediatric ophthalmology departments accounted for the maximum out-patient load compared to other sub-specialties like cornea, retina and glaucoma. Fellowship training was started in 3 hospitals and publications had come out from the research conducted by them. [14-27] The hospitals had publications on low vision services for children, refractive errors and ocular injuries in children. [14-17, 18,19, 21, 22] They tracked outcomes of pediatric cataract, strabismus and dacrocystorhinostomy surgeries. [20, 25, 27, 23]

The improved range of pediatric eye care service benefitted not just Nepal but neighboring Indian states of Uttar Pradesh, Bihar and West Bengal, which were the least developed parts of that country in terms of most socioeconomic indicators. [28] The International Centre for Eye Health, London and the Ministry of Health in Gambia, a small country in West Africa, had documented the reduction in the prevalence of blindness before and 10 years after the national eye care program. [9] The prevalence of blindness in children in a part of Nepal in 2015-16 was 0.07 % as compared to the prevalence of 0.14 % in 11-20 year olds in 1980. [4,29] The prevalence in UK for bilateral blindness was 6.3 per 10,000 children, that is 0.063% in 2007 [30] So the comprehensive effort over decades had almost halved the childhood blindness and it was now closer to prevalence in a developed country. The unilateral figures were higher due to trauma. [29,30]

The REACH program of Orbis India, aimed at tackling refractive errors in school children, was extended to Nepal in 2018. It standardized school screening and strengthened the partner hospital’s links with the community through the school eye program. [11] Better vision just does not make a child see better, it has implications in improving scholastic and sports performance and improving social interaction.

As Nepal prospers and develops, more preterm babies would be surviving, and retinopathy of prematurity would be there on the horizon, as it has been in neighbouring India. [31] The centres located in major urban areas, Kathmandu and Biratnagar, should liaison with neonatal intensive care units for screening of at-risk babies.

Training of anesthesiologists helped not just pediatric eye care but also pediatric orthopedic, otorhinolaryngology and general surgery. Though the turnover of the trained staff and relocation to other units was a concern, they work in other eye hospitals and continue to provide pediatric eye care services in the country. The interest for taking up pediatric ophthalmology as a separate sub-specialization was on the rise.

Doing pediatric eye surgeries is a bit like tiger conservation. Tiger is a symbol. For the tiger to be saved and their numbers to increase, an entire ecosystem has to be protected. Similarly, for every pediatric cataract identified, nearly 10,000 children have to be screened. This widespread screening of ocular and visual problems in the community upgrades and strengthens not just eye care, but also health care and educational systems. Better vision in children leads to academic betterment, more socialization and prevents disability. A better sighted child has more career options. The development and training of an eye care team would have far reaching consequences, not just for the pediatric ophthalmology department, but also for the entire hospital because the synergy created amongst doctors, paramedics, administrators and outreach team would have positive implications for efficient eye care service delivery. These experiences may be used for learning for other countries having population around 30 million.

All the hospitals showed a consistent increase in the volume of patient load and surgery performance of screening and surgical volume even after the end of the project. In addition to service delivery, many hospitals have stepped into clinical and operational research and were training mid-level ophthalmic personnel and medicos in children’s eye care.

## Data Availability

We have made available the data used for this manuscript in the form of an excel sheet

Legend 1/ Appendix 1: Data availability enclosed as Orbis data file.

## Notes

### Competing Interest Statement

The authors have declared no competing interest.

### Funding Statement

There was no specific funder for this work. It was done purely out of academic interest

### Author Declarations

Permission was sought and obtained from the ethics committee of Nepal Netra Jyoti Sangh, Kathmandu, Nepal. Annual reports of Orbis International, India Country Office, Nepal Netra Jyoti Sangh and Tilganga Eye Institute were studied as were the project reports, quarterly reports and midterm and final evaluation reports.

